# A Review on Calibration Methods of Cancer Simulation Models

**DOI:** 10.1101/2024.11.18.24317357

**Authors:** Yichi Zhang, Oguzhan Alagoz

## Abstract

Calibration, a critical step in the development of simulation models, involves adjusting unobservable parameters to ensure that the outcomes of the model closely align with observed target data. This process is particularly vital in cancer simulation models with a natural history component where direct data to inform natural history parameters are rarely available. This work reviews the literature of cancer simulation models with a natural history component and identifies the calibration approaches used in these models with respect to the following attributes: calibration target, goodness-of-fit (GOF) measure, parameter search algorithm, acceptance criteria, and stopping rules. After a comprehensive search of the PubMed database from 1981 to June 2023, 68 studies were included in the review. Nearly all (n=66) articles specified the calibration targets, and most articles (n=56) specified the parameter search algorithms they used, whereas goodness-of-fit metric (n=51) and acceptance criteria/stopping rule (n=45) were reported for fewer times. The most frequently used calibration targets were incidence, mortality, and prevalence, whose data sources primarily come from cancer registries and observational studies. The most used goodness-of-fit measure was weighted mean squared error. Random search has been the predominant method for parameter search, followed by grid search and Nelder-mead method. Machine learning-based algorithms, despite their fast advancement in the recent decade, has been underutilized in the cancer simulation models. More research is needed to compare different parameter search algorithms used for calibration.

**Key points:**

- This work reviewed the literature of cancer simulation models with a natural history component and identified the calibration approaches used in these models with respect to the following attributes: calibration target, goodness-of-fit (GOF) measure, parameter search algorithm, acceptance criteria, and stopping rules.
- Random search has been the predominant method for parameter search, followed by grid search and Nelder-mead method.
- Machine learning-based algorithms, despite their fast advancement in the recent decade, has been underutilized in the cancer simulation models. Furthermore, more research is needed to compare different parameter search algorithms used for calibration.

## 1 Introduction

Computer simulation models have been increasingly used to address cancer control problems. For example, the National Cancer Institute (NCI)’s Cancer Intervention Modeling and Surveillance Modeling Network (CISNET) simulation models have been used to inform the US Preventive Services Task Force screening recommendations for breast cancer ([1–3]), colorectal cancer [4], and lung cancer [5].

A crucial component of cancer simulation models is natural history, which represents the trajectory of cancer in an individual over time in the absence of a medical intervention. While a few of the natural history model parameters such as prevalence of cancer subtype could be estimated from primary data, most components such as the average tumor growth rate and the proportion of the tumors that regress remain unobservable. Consequently, in the absence of direct available data to estimate such parameters, models can determine the values of these parameters such that the model results match observed outcomes.

The most commonly used method to estimate directly unobservable parameters is *calibration*, which refers to the process of adjusting unobservable parameter values to ensure that the model’s outcomes align closely with observed target data such as observed incidence and mortality [6]. As contemporary cancer simulation models grow in complexity, resulting in a large number of natural history parameters that need to be estimated through calibration, and computational demands rise, modelers face the challenge of conducting efficient calibration. This requires an optimal compromise between parameter combinations that mirror clinical data and computational time and resource demands.

The simplest approach to implement calibration is to conduct a full-scale grid search for the entire parameter space, which involves discretizing the continuous parameters and running the simulation model for all possible combinations of the unobservable input parameters. While this is an easy-to-implement method, it requires major computational time due to the complexity of the models and a large number of input parameter combinations that need to be evaluated. For example, one study on calibrating an established breast cancer simulation model notes that a single replication of the model takes approximately 10 minutes on a stand-alone computer [7]. Considering that the study needed to evaluate approximately 400,000 input parameter combinations, the calibration procedure could take over 70 days to complete, which may not be computationally feasible. Not surprisingly, due to the need for speeding up the calibration in cancer simulation models, there is a growing interest in developing efficient strategies to search the parameter space for the calibration. A rich body of literature suggested using metaheuristic and structured methods such as grid search, random search, Nelder-Mead algorithm, neural networks, and Bayesian optimization for calibration [7–12].

Despite a drastic increase of interest in calibration from the cancer modeling community in the last decade, no study conducted a systematic review of the methods used for calibration in cancer simulation models, which is the focus of the present review. To our knowledge, only one previous study conducted a systematic review of the calibration methods used in the cancer simulation models and included studies published until 2006 [6]. That study did not focus on the optimization and heuristic methods used for the parameter search algorithm, instead, it primarily analyzed the studies based on modeled tumor types, metrics used for measuring goodness-of-fit of the models, and validation strategies. Compared to that study, we included more recent studies, which is crucial since there has been a major increase in the number of studies utilizing calibration methods and the diversity of the calibration approaches. In addition, our emphasis in this work is to classify the studies with respect to the calibration methods used, which provides insight into the preferred calibration methods by modelers in cancer research.

## 2 Methods

### 2.1 Search Strategy

We conducted a systematic review on calibration methods employed in cancer simulation models with a natural history component. A comprehensive search of the PubMed database from 1981 to June 2023 was performed for articles published in English. Our search criteria only considered papers that contain “simulation” and “cancer” in title/abstract and “calibration” in the main text. Note that PubMed automatically includes synonymous or related keywords. For instance, in the case of ‘cancer’, PubMed also identifies words like ‘neoplasm’ or ‘cancers.’ In situations where multiple papers were published based on the same study, only one was included in our review. Additional studies were found through manual searches of the studies that cited tutorial papers describing simulation calibration and the previous systematic review article.

Our inclusion criteria consisted of articles that developed a cancer simulation model with a natural history component and calibrated the natural history component to match specific calibration targets. We excluded preprint/nonrefereed articles as well as articles focusing on models of disease other than cancer, with the exception of Human Papillomavirus due to its profound association with cervical cancer. We also excluded models devoid of a natural history component or those that do not calibrate their parameters. Tutorial articles that describe specific calibration methods were also excluded.

### 2.2 Classification of the Studies and Reporting of the Results

Our search focused on the following attributes of the calibration articles: calibration target, goodness-of-fit measure, parameter search algorithm, acceptance criterion and stopping rule. We briefly describe them here.

*Calibration targets* are observed empirical data, such as cancer incidence and mortality rates, that can be directly estimated. As the name implies, these targets serve as benchmarks that a model aims to replicate during the calibration process. Typical calibration targets include incidence (the rate of new cancer cases within a specific period), mortality (the death rate due to cancer), survival (the proportion of patients living for a certain time after diagnosis), and stage distribution (the breakdown of cancer cases by cancer stage at diagnosis). These critical data are sourced from cancer registries, which collect comprehensive cancer patient information; observational studies, which monitor subjects in natural conditions without intervention; and randomized controlled trials, where subjects are randomly assigned to experimental or control groups to test the efficacy of treatments.

*Goodness-of-fit* (GOF) of a simulation model reflects how well the results of the running the model using an input parameter combination align with calibration targets. A straightforward GOF measure involves visual comparison where model outputs are manually contrasted with calibration targets to evaluate their fit, which is not ideal due to the subjectivity, hence, most models employed quantitative measures. Predominantly used GOF measures are mean squared error (MSE), weighted mean squared error, likelihood, and confidence interval envelope. Choosing a GOF measure that is compatible with simulation model is crucial for the model’s success. We provide a formal description for each GOF metric used in the articles included in our study in **Appendix A**.

*Parameter search algorithms* refer to the methods to identify parameter combinations that are sufficiently close to the calibration targets. The degree of closeness between parameter combinations and calibration targets is measured by the GOF measure, serving as an error function in the parameter search algorithms. This problem can be conceptualized as finding the parameter combination that minimizes the GOF measure across the parameter space, a topic that has been comprehensively studied. Additionally, the parameter space is typically expansive and non-convex due to the nonlinear nature of the simulation model or constraints on its parameters, rendering it challenging to find a global optimum using an algorithm with a reasonable runtime. Despite this, a selection of alternative heuristic algorithms can be utilized to find parameter combinations that reasonably approximate the calibration targets within an acceptable runtime. We describe each parameter search algorithm used in the articles included in our study in **Appendix B**.

*Acceptance criteria* refer to the standards set by the modelers to determine if predictions made by the model using a particular input parameter combination align sufficiently well with the calibration targets. These criteria are typically measured using the GOF metrics. On the other hand, *stopping rules* refer to the thresholds or conditions that, once met, lead modelers to terminate the calibration process. Common stopping rules include identifying an adequate number of parameters that fulfill the acceptance criteria or reaching a pre-specified number of iterations/time during calibration.

## 3 Results

### 3.1 Search results

Our search resulted in 253 unique articles. Of these, 56 met the inclusion criteria, while the remainder were excluded for the reasons mentioned previously. An examination of the references within these articles led us to identify 12 additional articles that satisfied our criteria, bringing the total to 68 articles.

Prior to 2006, very few articles met our inclusion criteria. A notable uptick in articles that fit our criteria began after 2006, peaking at 8 articles per year in 2019 (**Figure 1**). Breast, colorectal, and cervical were the predominant cancer types represented with these simulation models, with 13 (19.1%), 13 (19.1%), and 11 (16.2%) articles respectively, as shown in **Table 3**. This contrasts with the less common cancer types, such as skin or thyroid cancers, which were each modeled in only a single article.

**Figure 1.**
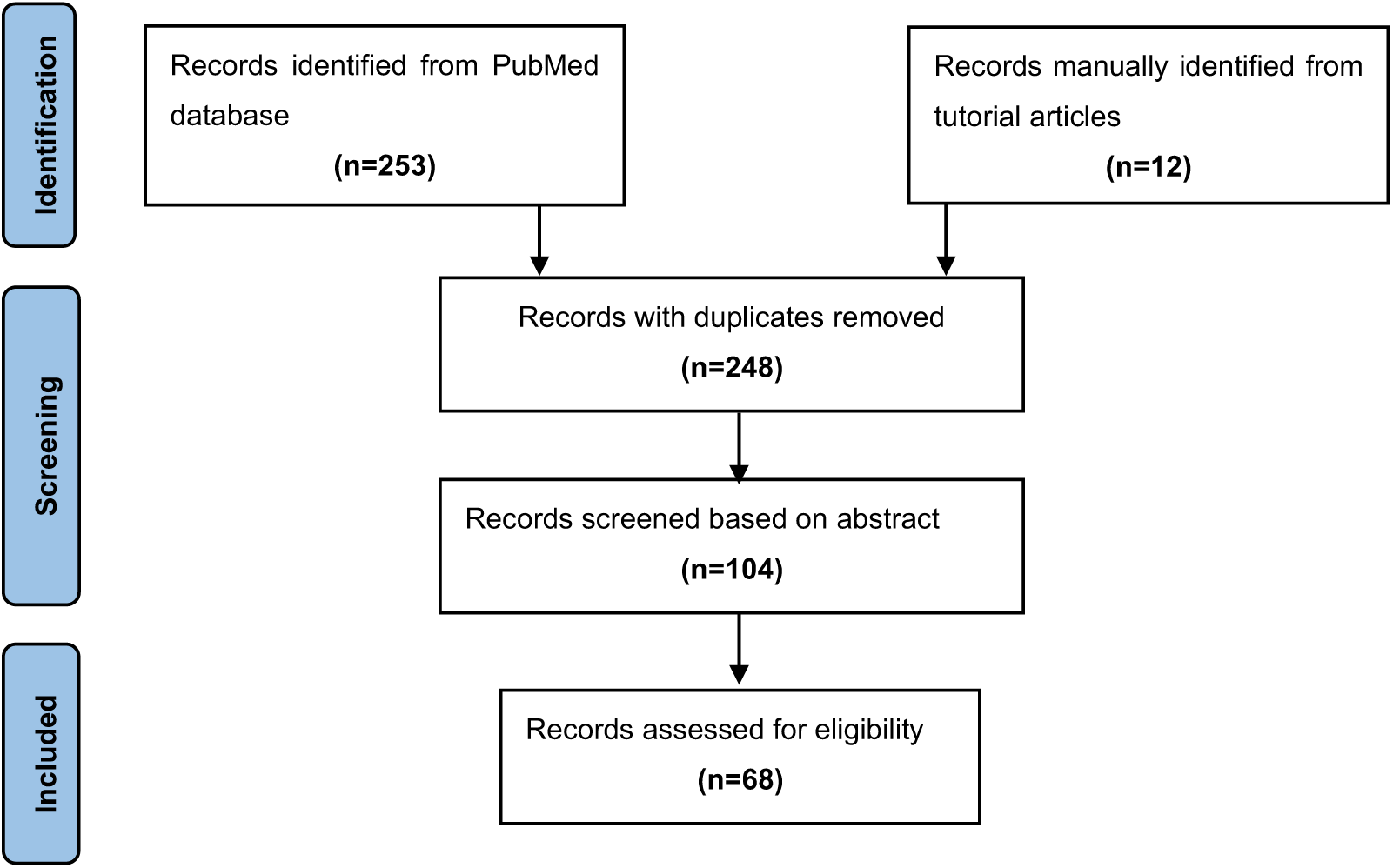
PRISMA flow diagram for search strategy.

**Figure 2.**
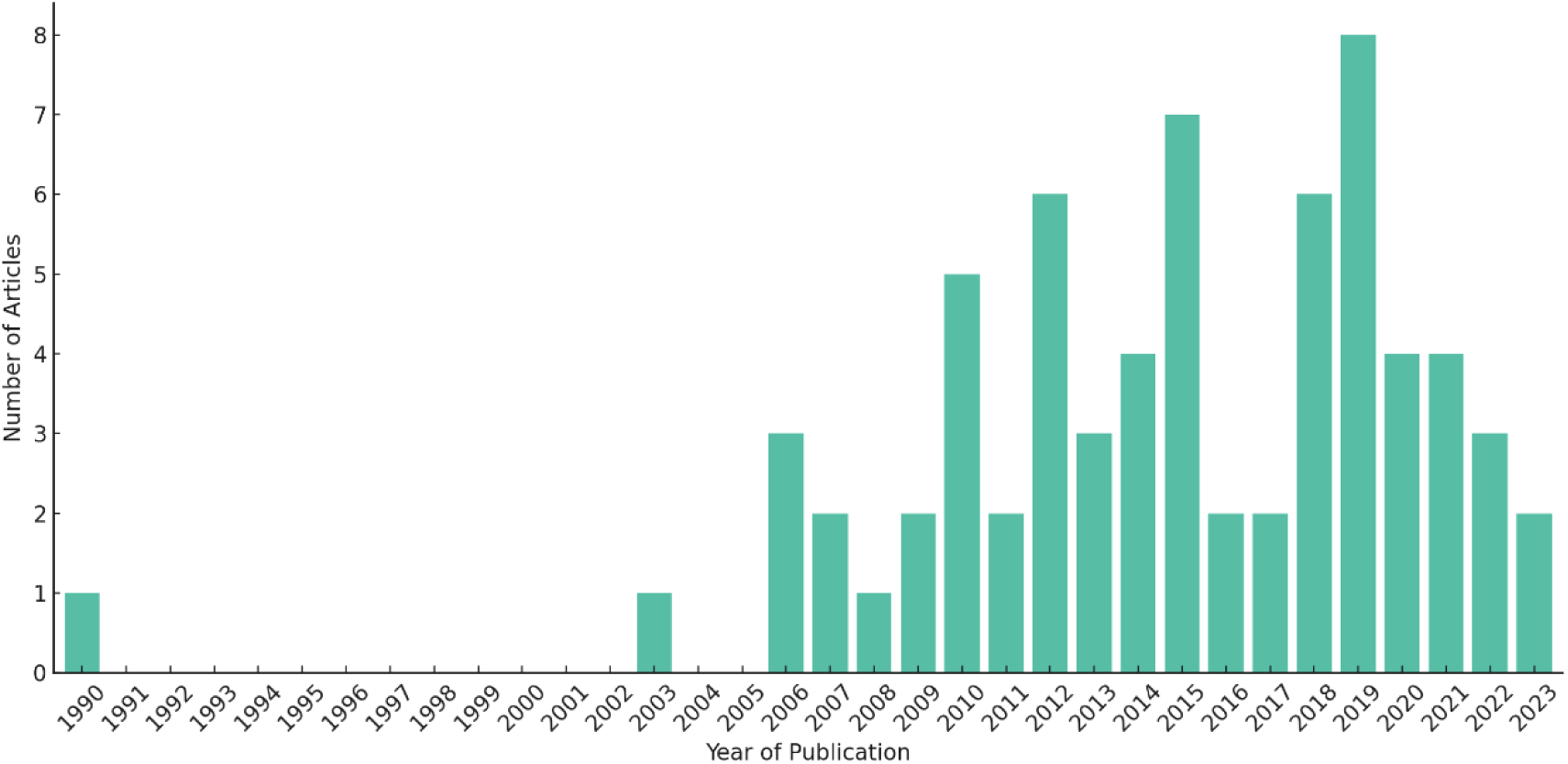
Number of cancer simulation models that utilized calibration by the year of publication.

In our literature review, 66 (97.1%) of the 68 articles mentioned their calibration targets. Among these, one article did not specify the data source for targets, and three articles did not specify the target type. The most commonly used calibration data source is the cancer registry, which was used by 43 (63.2%) articles (**Table 1)**, notably the United States National Cancer Institute’s Surveillance, Epidemiology, and End Results (SEER) program. This is followed by observational studies, which include a variety of subcategories, including cohort studies, retrospective studies, and national surveys [13–15]. Thirteen (19.1%) articles utilized data from randomized controlled trials. The most frequently used target types are incidence, mortality, and prevalence, respectively (**Table 2**). Only 19 (28.0%) of the 68 articles used a single type of calibration target type.

**Table 1.** Data source types for the calibration targets used in the cancer simulation models.

| Data source type | Number of studies | Reference |
| --- | --- | --- |
| Cancer registry | 42 | [8], [10], [18], [12], [19], [20], [21], [22], [23], [24], [25], [14], [13], [26], [7], [27], [11], [28], [29], [30], [31], [32], [33], [34], [35], [36], [37], [38], [39], [40], [41], [42], [43], [44], [45], [46], [47], [48], [49], [50], [51], [52] |
| Observational study | 24 | [9], [8], [10], [18], [12], [24], [14], [13], [26], [53], [54], [29], [55], [35], [36], [56], [57], [40], [41], [58], [59], [48], [60], [61] |
| Randomized controlled trial | 13 | [62], [16], [63], [64], [65], [66], [34], [56], [67], [68], [46], [59], [69] |
| Other | 1 | [70] |
| Unspecified | 3 | [71], [72], [73] |

**Table 2.** Calibration targets used in the cancer simulation models.

| **Calibration target** | **Number of studies** | **Reference** |
| --- | --- | --- |
| Incidence | 36 | [62], [9], [12], [63], [64], [20], [22], [25], [13], [71], [7], [27], [11], [54], [30], [32], [55], [34], [35], [36], [37], [38], [39], [67], [40], [41], [43], [44], [45], [46], [58], [47], [48], [60], [49], [52] |
| Mortality | 18 | [9], [12], [63], [19], [20], [23], [14], [27], [28], [33], [36], [37], [40], [41], [42], [68], [45], [60] |
| Prevalence | 18 | [18], [12], [24], [14], [26], [71], [53], [11], [29], [31], [55], [34], [35], [67], [41], [42], [48], [69] |
| Survival | 8 | [21], [25], [13], [65], [66], [37], [56], [48] |
| Stage distribution | 5 | [64], [30], [36], [38], [42] |
| Other | 4 | [70], [57], [59], [61] |
| Unspecified | 5 | [8], [72], [73], [50], [51] |

**Table 3.** Cancer types represented in the cancer simulation models.

| **Cancer type** | **Number of studies** | **Reference** |
| --- | --- | --- |
| Breast | 13 | [63], [19], [20], [22], [65], [7], [27], [38], [39], [73], [40], [44], [45] |
| Colorectal | 13 | [8], [18], [24], [26], [71], [66], [30], [37], [70], [57], [74], [59], [48] |
| Cervical | 11 | [10], [12], [53], [54], [31], [35], [67], [72], [58], [69], [50] |
| Lung | 9 | [62], [16], [14], [36], [46], [60], [51], [52], [61] |
| Prostate | 7 | [64], [25], [34], [56], [68], [75], [49] |
| Multiple | 4 | [23], [13], [28], [33] |
| Esophageal | 3 | [11], [55], [47] |
| Pancreatic | 2 | [41], [42] |
| Soft tissue | 1 | [9] |
| Urologic | 1 | [21] |
| Anal | 1 | [29] |
| Thyroid | 1 | [32] |
| Skin | 1 | [43] |
| Bladder | 1 | [15] |

Of the 68 articles, the weighted MSE was the most employed GOF metric (**Table 4**). Specifically, 7 (10.3%) articles utilized chi-squared error as their weight, and 10 (14.7%) articles incorporated other types of weights. Following the weighted MSE, the MSE without weights was the second most popular metric, being used in 15 (22.1%) articles. Likelihood was the third most frequent GOF metric, with 10 (14.7%) articles using it. 17 (25%) of the articles do not specify their choice of GOF metrics.

**Table 4.** Goodness-of-fitness measures used in the cancer simulation models.

| **Goodness-of-fitness measure** | **Total number** | **Reference** |
| --- | --- | --- |
| Sum of square | 15 | [16], [18], [20], [21], [23], [24], [66], [28], [33], [36], [44], [68], [74], [60], [61] |
| Likelihood | 10 | [10], [71], [53], [29], [31], [35], [40], [41], [42], [58] |
| Weighted sum of square | 10 | [12], [13], [26], [30], [56], [39], [67], [45], [46], [15] |
| Chi square (weighted sum) | 7 | [63], [64], [11], [32], [55], [38], [47] |
| Envelope method | 3 | [19], [7], [27] |
| Other | 7 | [26], [65], [34], [72], [43], [51], [52] |
| Unspecified | 17 | [62], [9], [8], [22], [25], [14], [54], [37], [70], [57], [73], [75], [59], [48], [69], [49], [50] |

**Table 5.**
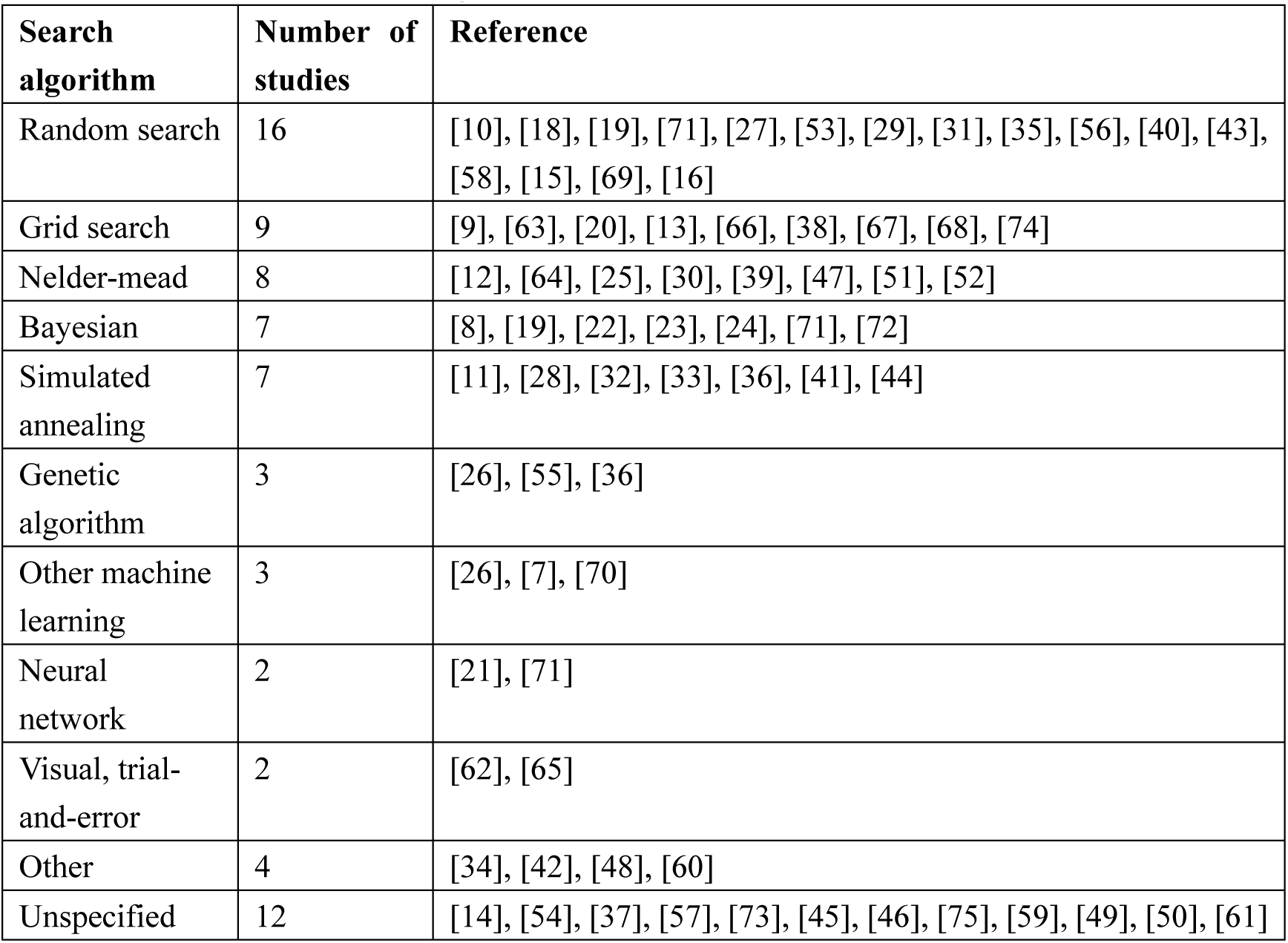
Parameter search algorithms used in the cancer simulation models.

| **Search algorithm** | **Number of studies** | **Reference** |
| --- | --- | --- |
| Random search | 16 | [10], [18], [19], [71], [27], [53], [29], [31], [35], [56], [40], [43], [58], [15], [69], [16] |
| Grid search | 9 | [9], [63], [20], [13], [66], [38], [67], [68], [74] |
| Nelder-mead | 8 | [12], [64], [25], [30], [39], [47], [51], [52] |
| Bayesian | 7 | [8], [19], [22], [23], [24], [71], [72] |
| Simulated annealing | 7 | [11], [28], [32], [33], [36], [41], [44] |
| Genetic algorithm | 3 | [26], [55], [36] |
| Other machine learning | 3 | [26], [7], [70] |
| Neural network | 2 | [21], [71] |
| Visual, trial-and-error | 2 | [62], [65] |
| Other | 4 | [34], [42], [48], [60] |
| Unspecified | 12 | [14], [54], [37], [57], [73], [45], [46], [75], [59], [49], [50], [61] |

**Table 6.** Acceptance criteria and stopping rule used in the cancer simulation models.

| **Acceptance criteria/stopping rule** | **Number of studies** | **Reference** |
| --- | --- | --- |
| Neither acceptance criteria not stopping rule is used | 21 | [16], [64], [22], [7], [27], [11], [28], [29], [30], [31], [33], [38], [39], [41], [42], [43], [68], [58], [74], [47], [61] |
| Both acceptance criteria and stopping rule were used | 14 | [9], [10], [23], [24], [26], [53], [66], [32], [55], [35], [36], [67], [72], [40] |
| Only stopping rule was used | 6 | [18], [12], [71], [65], [37], [69] |
| Only acceptable criteria was used | 3 | [19], [21], [44] |
| Unspecified | 23 | [62], [8], [63], [20], [25], [14], [13], [54], [34], [56], [70], [57], [73], [45], [46], [75], [59], [69], [60], [49], [50], [51], [52] |

Most articles we found in the literature search describes their parameter search algorithms. Of the 68 included articles, random search emerged as the most favored parameter search algorithm, being referenced in 16 (23.5%) articles. Note that the specific usage of the random search method is occasionally inferred rather than explicitly stated. For example, Hammer et al. (2019) [16] describes their parameter search algorithm as “the simulation was run 10,000 times, with each run using a number randomly drawn from the range of possible growth rates.” Such descriptions hint at the possible use of random search, but also raise questions about the exact methodology applied since other more complicated algorithms such as simulated annealing also randomly draw the number from plausible range. It is possible that some models might be leveraging more nuanced algorithms but might not be detailing them adequately in their documentation.

Following random search in popularity are the grid search and Nelder-mead methods, each being adopted in 9 (13.2%) and 8 (11.8%) articles respectively. Interestingly, despite the rising interest in machine learning (ML) for analyzing large datasets, as is typical with simulation models, only five articles utilized ML-based strategies. Furthermore, 12 (17.6%) articles did not specify their calibration algorithm, often leaving other calibration details ambiguous as well.

In our search, 21 (30.8%) articles reported to have terminated the calibration process after evaluating a designated number of combinations. These articles then selected the top-performing combinations without explicitly stating their acceptance criteria. A total of 14 (20.6%) articles clearly specified both acceptance criteria and stopping rule. Six and three articles only specify stopping rule and acceptance criteria, respectively. A total of 23 (33.8%) articles did not provide any information on acceptance criteria or stopping rule.

## 4 Discussion

Calibration of natural history parameters in cancer simulation models is typically the most time-consuming component of building a model, making a careful selection of efficient parameter search algorithms necessary. In addition, the choice of suitable GOF metrics, acceptance criteria, and stopping rule with the algorithm is also crucial for calibration. Our review aims to summarize the strategies adopted by modelers in selecting these essential components, providing insights into current practices and potential improvements in simulation model calibration.

MSE, in both weighted and unweighted forms, is the most commonly used GOF measure, and it is considered as the default method in many implementations. However, prior to finalizing the choice of a GOF measure for a model, it is important to conduct a thorough comparison among various GOF measures. This is crucial because each measure has its unique strengths and limitations, and its performance varies in different scenario. For instance, MSE is sensitive to outliers, leading to a heavy penalty for larger errors. Secondly, MSE may not allow the capturing the temporal trends in observed outcomes such as incidence, which may be significant for accurate representation. For example, a recent thyroid cancer simulation model reported that successful replication of thyroid cancer epidemiology in the US requires modeling the drastic increase in thyroid cancer incidence between 1990s and 2010s, and the use of MSE may not allow choosing parameters combinations to reflect this temporal trend [17].

Given the advancement of machine learning algorithms in the recent decade, it is surprising that only a small number of studies, specifically five articles in our review, have utilized machine learning-based algorithms. This observation suggests a potential underutilization of these powerful tools in the field. Modelers and researchers should be aware that machine learning is highly accessible. In-depth expertise in machine learning is no longer a prerequisite for implementation, thanks to the proliferation of user-friendly libraries and frameworks. These resources efficiently manage the intricate aspects of algorithmic processing, so the users only need to know coding to implement such algorithms. However, users still need machine learning knowledge to pick the suitable algorithms specific to the model.

We identified only one prior systematic review that conducted systematic reviews of calibration process [6]. This review, however, is limited to articles published before 2006 and does not concentrate on parameter search algorithms. Only a small proportion of the articles they included have clearly documented their search algorithm during calibration process, therefore the comparison of our present study’s findings to that article is not possible.

Our review also highlighted the need for reporting of the calibration methods in simulation modeling papers. Many of the studies included in this review lack a comprehensive description of their calibration process, including how they selected the GOF metric and parameter search algorithm, as well as the absence of acceptance criteria and stopping rules. To enhance transparency and understanding, we advise authors to include a detailed account of the calibration procedure in the main body of the text or in a supplement. This addition would significantly help readers in understanding the full extent and robustness of the calibration process.

In addition to providing a detailed account of the calibration procedure, we strongly encourage modelers to consider the possibility of making their source code publicly available, given that the nature of their project permits it. Future researchers who are interested in the model can gain deeper understanding of the model and can potentially build upon the current model.

### 4.1 Future Research Directions for Calibration of Simulation Models

The strong need for computationally efficient simulation calibration methods and growing interest from the cancer research community on simulation calibration provide an opportunity for future research. There is a noticeable lack of articles that perform a comparative analysis of multiple parameter search algorithms used for calibration. This omission results in ambiguity regarding the selection of the most suitable parameter algorithms. A more thorough comparison in future studies could provide valuable insights into the efficacy and applicability of different algorithms. A comparison would also make the calibration process more robust. Furthermore, considering the significant advancements in machine learning, its current application in the field appears underutilized. We encourage more studies to explore the incorporation of machine learning-based algorithms, especially given the ease of implementation afforded by contemporary libraries. The integration of these advanced techniques could lead to more refined and efficient modeling approaches.

### 4.2 Limitations

Our study has several limitations. First, while numerous parameter search algorithms are categorized under random search, making it the most commonly used algorithm, this classification may be misleading due to a lack of detailed information. In cases where only the use of randomness is mentioned without further details, the classification may not accurately reflect the actual algorithm used in the model. Secondly, it is possible that some authors utilized multiple parameter search algorithms in their papers but only reported the most successful one in the paper and omitted the details for others. Similarly, studies may have used a combination of search algorithms.

## 5 Conclusion

This study summarizes the calibration methods used by cancer simulation models. We found that the most common used parameter search algorithm is random search and the most commonly used GOF metric is MSE. Given the recent advancement of machine learning techniques, we found fewer than expected number of models adopting this method. The findings also signal a critical need for enhanced transparency and standardization in reporting calibration processes. Detailed documentation of the calibration methods is essential for replicability and further methodological advancements.

## Data Availability

not applicable

## Notes

**Funding source:** This work was supported by the National Institutes of Health (NIH) under National Cancer Institute Grant R01CA251566. The funding agreement ensured the authors’ independence in designing the study, interpreting the data, writing, and publishing the report.

**Conflict of Interest statement**: Dr. Alagoz reports grants from NIH, during the conduct of the study; personal fees from Bristol Myers Squibb, personal fees from Exact Sciences, other from Innovo Analytics, outside the submitted work. Mr. Zhang has no conflict of interest to report.

### Competing Interest Statement

Dr. Alagoz reports grants from NIH, during the conduct of the study; personal fees from Bristol Myers Squibb, personal fees from Exact Sciences, other from Innovo Analytics, outside the submitted work. Mr. Zhang has no conflict of interest to report.

### Funding Statement

This work was supported by the National Institutes of Health (NIH) under National Cancer Institute Grant R01CA251566. The funding agreement ensured the authors independence in designing the study, interpreting the data, writing, and publishing the report.

