## Supplement for "A Review on Calibration Methods of Cancer Simulation Models"

### APPENDIX

### Appendix A. Details of the goodness-of-fitness measures used in the articles included in this review

#### A.1. Mean squared error

The mean squared error (MSE) is a commonly used and intuitive goodness-of-fit (GOF) measure. MSE quantifies the average of the squared differences, referred to as errors, between the model output derived from a specific parameter combination and the corresponding calibration target. The formula for MSE is expressed as:

$$\text{MSE} = \frac{1}{n}\sum_{i=1}^{n}\left(Y_i - \widehat{Y}_i\right)^2$$

In this formula, $Y_i$ and $\widehat{Y}_i$ represent the $i^{\text{th}}$ calibration target and the $i^{\text{th}}$ model output of a specific parameter combination, respectively. MSE can be interpreted as the distance between the calibration targets and the model output since it calculates the average of the square of Euclidean distances. As such, MSE is always non-negative (reaches zero when the parameter combination coincides with the calibration targets), with larger values indicative of poorer fit. However, a key limitation of MSE is its susceptibility to extreme values as stated in the main text.

#### A.2. Weighted mean squared error

This is similar to MSE, but weighted MSE multiplies each square of the error by a weight ($w_i$).

$$MSE = \frac{1}{n}\sum_{i=1}^{n}\left(w_i\left(Y_i - \widehat{Y}_i\right)^2\right)$$

The weight can be different across the calibration targets and reflects our prior knowledge on the calibration targets. For instance, to mitigate MSE's poor performance with extreme values, we can set wi=1/ Yi. In this case, if the value of one calibration target significantly outstrips others, its error, now tempered by weights, is less likely to excessively influence the weighted MSE. When setting to 1/Yi, it is also the Pearson's chi-squared measure, one of the most popular weighted mean squared error. Furthermore, weights can be leveraged to signify our preference for certain calibration targets, by assigning greater weight to those deemed more critical.

##### A.3. Likelihood

The likelihood measure is derived from maximum likelihood estimation (MLE). The goal of MLE is to identify the parameter set that makes the calibration targets most probable. In other words, MLE aims to maximize the likelihood that a model will yield calibration targets. The likelihood measures the joint probability of producing the entire set of calibration targets, denoted by $a$, given a set of parameters, denoted by $\theta$, and the formula is given below:

$$L(\theta) = \prod_{a} P(a|\theta)$$

However, there is an inherent challenge with the likelihood measure when dealing with calibration targets with many entries. As each $P(a|\theta)$ is a fractional value between 0 and 1, multiplying numerous such probabilities results in a product that can be extremely close to zero. This makes direct comparisons of likelihood values for different parameter combinations difficult due to their minuteness. To circumvent this, researchers often take the logarithm of the likelihood. This transformation, which converts the products into summations of log probabilities, not only makes the computations more manageable but also avoids the dilemma of vanishingly small likelihood values.

Additionally, there's another nuance to be mindful of. Typically, in optimization scenarios, one seeks to minimize an error function. However, with raw likelihoods, a higher value signifies a better fit, which goes against the conventional optimization logic. To standardize the measure with other error metrics like MSE, where lower values are preferred, researchers work with the negative logarithm of the likelihood. Negative logarithm of the likelihood measures the unlikeliness to produce the calibration targets given the current parameter combinations, which ensures that lower values of the negative log-likelihood signify better model fits, aligning with the general optimization paradigm.

##### A.4. Confidence interval score

This measure is particularly useful when confidence intervals have been computed for the calibration targets. The confidence interval score is determined by counting the number of model outputs that fall out of the corresponding confidence interval of the calibration targets, with a smaller score indicating a superior fit of the parameter combination. Additionally, weights can be assigned to the confidence interval score to reflect varying levels of significance. Depending on the degree of accuracy required, a wider or narrower confidence interval can be used.

### APPENDIX B. Details of the parameter search algorithms used in the articles included in this review

#### B.1. Grid search

The grid search method starts with identifying a parameter space that encompasses all feasible parameter combinations. For each calibrated parameter, a plausible range is determined such that a parameter combination is deemed feasible if each calibrated parameter within the combination resides within the plausible range. Subsequently, all plausible ranges are partitioned into distinct segments, giving rise to the concept of a "grid." The grid search method executes the simulation model with each parameter combination in the grid. For instance, if there are five calibrated parameters, and each parameter's range is divided into three segments, the grid search will run the model $3^5$ (243) times, seeking the parameter combination with the minimum GOF measure. However, the primary drawback of this method is its computational intensity. For a model with 30 calibrated parameters, even if we only bisect the parameter range, we would need to run our model approximately $2^{30}$ times, which is roughly equivalent to a hundred million iterations. As such, grid search is typically recommended only for models with a limited parameter space. For instance, in the model by [1], only four parameters required calibration, making grid search a suitable choice. Similarly, [2] utilized grid search for neural network hyperparameters, which represented a significantly smaller challenge compared to the model's overall complexity.

#### B.2. Random search

The random search method has been the most prevalent method for parameter searching. In this approach, appropriate distributions for all input parameters are established, either based on expert opinions or through literature reviews. Often, researchers specify a "plausible range," which can be interpreted as a uniform distribution where minimum and maximum value of the distribution is the lower and upper bound of the range, respectively. Parameter combinations are subsequently sampled from these pre-established distributions, and the GOF measures for these combinations are determined by executing the model. This procedure continues until either a set maximum number of runs is reached or enough parameter combinations yielding a satisfactory GOF measure is identified.

The random search method is an intuitive algorithm that is simple to implement. However, it lacks a strategic direction in guiding the search, which can result in inefficiencies, particularly in situations with a large parameter space. The inherent randomness may cause uneven exploration, with some regions oversampled and others left untouched. Thus, the method doesn't ensure convergence to a global optimum within a computationally feasible timeframe.

To address the shortcoming, several refined variants of the random search method have been proposed. One example is Latin Hypercube Sampling (LHS) method. Instead of relying on pure randomness, LHS ensures more consistent sampling across the parameter space. Initially, one determines the number of sampling combinations. Subsequently, the feasible distribution of all parameters into that many subranges. For each parameter combination, the values are randomly drawn from these subranges, and each subrange is used only once. This ensures a more systematic coverage of the parameter space, making LHS more efficient in terms of exploration compared to plain random search. We suggest against using plain random search algorithm because variants like LHS have improved performance without any major drawbacks. Modelers should select variants based on the model's specific needs.

##### B.3. Simulated annealing

Simulated Annealing (SA) ranks as the fourth most popular algorithm in our search, being employed in seven papers. Drawing its inspiration from the metallurgical annealing process, SA mimics how metals undergo controlled cooling to alter their properties. Within the algorithmic context, a metaphorical 'temperature' governs the probability of accepting parameter combinations with poorer performance, allowing the search to potentially escape local optima. The temperature parameter in the SA approach is high initially, introducing greater randomness and flexibility in the search. As the algorithm progresses, this temperature gradually decreases, tightening the acceptance criterion and becoming more deterministic in its choices.

For SA to function effectively, certain foundational elements must be defined. One of these is the notion of 'neighborhood' for a given point in the parameter space. For instance, a neighbor might be defined by constraining the Euclidean distance between the current point and its neighboring candidates. The way neighbors are defined is crucial as it influences the search dynamics and, consequently, the algorithm's performance. Additionally, the algorithm relies on efficient techniques to sample or select random neighbors. We also need to find a good temperature function that controls the acceptance of neighbors with worse fit, usually through trial and error because this is dependent on local extremum and thus model specific.

Formally, the SA algorithm operates as follows: Initially, a random point in the parameter space is chosen as the 'current' point and the temperature is set to a high value. For each subsequent iteration, a random neighbor of the current point is selected. If this neighboring point offers an improvement over the current point, it is immediately accepted. However, if we only accept better point, it is likely to be trapped in local extremum, and the real magic of SA lies in its ability to accept worse points based on a probability which is influenced by the current temperature. This probabilistic acceptance mechanism provides the algorithm the flexibility to escape from local optima traps, especially during the early stages when the temperature is high. As iterations progress and the temperature drops, the likelihood of accepting inferior parameter combinations diminishes, allowing the algorithm to stay in on an optimal or near-optimal combinations.

##### B.4. Nelder-Mead algorithm

Nelder-Mead algorithm ranks as the third most popular algorithm in our search, being used in eight papers. This algorithm operates by maintaining a simplex, a polytope consisting of n+1 parameter combinations within an n-dimensional parameter space. During each iteration, the algorithm tries to refine the worst-fitting parameter combination, using a series of operations such as reflection, expansion, contraction, and shrinkage. A standout advantage of the Nelder-Mead algorithm is its ability to optimize even when the objective function isn't differentiable. Nevertheless, it isn't without drawbacks. The algorithm is known for its slow convergence, demanding more computational resources. Moreover, the initial construction of the simplex plays a crucial role. If not chosen cautiously, there's a risk of the model either converging extremely slowly or becoming trapped at a local extremum.

##### B.5. Genetic algorithm

The Genetic Algorithm (GA) is a less commonly adopted approach, with only three papers in our study utilizing it. Drawing inspiration from natural selection and genetic principles, GA starts with a randomly generated cohort of feasible parameter combinations, or "chromosomes." This iterative algorithm progresses through successive "generations" to hone the fit of these combinations. In each generation, chromosomes are selected based on their fitness to the calibration targets, with better-performing combinations more likely to be chosen for reproduction. The reproductive mechanism, termed "crossover," emulates biological reproduction: selected parent chromosomes exchange data segments to birth new offspring chromosomes. To ensure genetic diversity and prevent convergence to local extremum, random mutations are introduced to the offspring. After the offspring are generated, some of the older generation chromosomes are replaced by these offspring. This iteration of selection, crossover, mutation, and replacement is repeated until a set termination criteria is reached, be it a specific level of GOF measure or a predetermined number of iterations. Over time, the algorithm promotes the survival of the fittest parameter combinations, resulting in a cohort that has evolved towards better fits to calibration targets.

Implementing the Genetic Algorithm (GA) requires meticulous attention to various details. One pivotal decision is determining the size of the cohort. A large cohort may escalate computational complexity beyond manageability, while a diminutive one risks diminishing the effectiveness of the GA. Selection functions, which determine the chromosomes chosen for reproduction, are equally crucial. Modelers must strike a balance between strategies like "elitist selection," which only allows the best-fit chromosomes for reproduction, and more randomized selection approaches that ensure diversity and allow confinement to local extremum. Modelers must decide on the number of parents involved in each crossover and set probabilities for crossover and mutation events. Beyond these primary genetic operators, there exist lesser-known operators, such as regrouping. These myriad decisions collectively shape the efficiency and outcomes of the GA process.

The main drawbacks of GA are that it is not as efficient as other algorithms with large parameter space. It is also easier to be trapped in local extremum.[3] found that SA outperformed GA in their Lung Cancer Policy Model.

#### B.6. Neural network

Neural networks have gained increasing popularity in the field of simulation modeling, partly attributable to their proficiency in accurately managing models with large input sizes. These networks bear similarity to the human brain, with artificial neurons mirroring biological brain cells and inter-neuron connections reflecting the signal transmission between neurons. Neurons are grouped into layers, consisting of an input layer, an output layer, and potentially multiple hidden layers. For each neuron beyond the input layer, its value is dictated by the following formula:

$$w\_\{ij\} = \sigma(\sum_{m=1}^{n} e_{mj} w\_\{i-1m\})$$

Here, $e_{mj}$ represents the weight of the edge set for optimization, and σ is the activation function that introduces nonlinearity into the neural network. In the absence of nonlinearity, the neural network reverts to predicting linearly separable cases. Frequently employed activation functions include the Rectified Linear Unit (ReLU) and the sigmoid function.

#### B.7. Bayesian method

Bayesian method originates from Bayes' theorem, $P(\theta|D) = P(D|\theta)P(\theta)/P(D)$

where $P(D|\theta), P(\theta|D), P(D), P(\theta)$ denotes the likelihood, the posterior, the evidence, and the prior. Since P(D) is fixed, the essence of the Bayesian method is the prior $P(\theta)$ — which encapsulates our pre-existing knowledge of the model — and the likelihood $P(D|\theta)$, reflecting how close our model conforms to the data. Together, they characterize the posterior $P(D|\theta)$, which represents the revised understanding of the parameters $\theta$ based on the data.

In our search, Bayesian method ranked the third most used algorithm with 9 papers using it. We found its influence extends far beyond papers that solely focus on Bayesian analysis. The principles of Bayesian reasoning are embedded in various other methodologies, even if they're not labeled as "Bayesian". For example, [4] uses random search with likelihood score as its GOF metrics. The likelihood score, representing the fit of a model to the data, can be analogized to the Bayesian concept of updating beliefs about model parameters given new data. Moreover, Bayesian method's emphasis on distributions (instead of single-point estimates) aligns well with contemporary modeling challenges. In complex systems, it's often more informative to understand the range and uncertainty of possible outcomes, either characterized by a probability distribution or by multiple parameter combinations rather than fixating on a single optimal solution.
